# Radiofrequency Catheter Ablation of Atrial Fibrillation in Patients with Anomalous Pulmonary Venous Return: A Multicenter Case Series

**DOI:** 10.64898/2026.09.30.26363976

**Authors:** Camilo A. Estrada-Vargas, Omar J. Baqal, Mahiraj Singh Sidhu, James Y. Kim, Vinicius de Sousa Barzon Serra, Aakash Somappa, Juan F. Rodriguez-Riascos, Carlos A. Rojas, Komandoor Srivathsan

## Abstract

**Background:** Atrial fibrillation (AF) has been observed in adults with anomalous pulmonary venous return (APVR), but data on catheter ablation outcomes in this population remain limited to case reports and small series.

**Methods:** We retrospectively reviewed 14 adult patients with confirmed APVR who underwent catheter ablation for AF at three tertiary academic centers in the United States between July 2003 and May 2025. Baseline, procedural, electroanatomic mapping, and outcome data were collected. Freedom from atrial arrhythmia recurrence was estimated by Kaplan-Meier analysis.

**Results:** Mean age was 56 ± 13 years and 79% were male. Heart failure was present in 64%, and hypertension and coronary artery disease in 43% each. Most had partial APVR (86%), and 57% had prior surgical correction. Radiofrequency ablation was used in all patients, combined with pulsed-field ablation in one. Acute success was achieved in all patients, although areas with residual pulmonary vein or SVC electrical activity were left untreated in 3 (21%). Non-pulmonary-vein electrical activity was more common in patients with prior surgical correction than in those without (43% vs 17%). Median follow-up was 3.4 years (IQR, 2.0-9.6). Freedom from arrhythmia recurrence was 78.6% at 1 year and 53.9% at 5 years. Two patients (14%) required reablation; complications occurred in 3 (21%) without long-term sequelae.

**Conclusions:** Catheter ablation was acutely successful in all patients in this largest reported APVR cohort, despite substantial comorbidity burden and anatomic complexity. The arrhythmogenic substrate often extended beyond the anomalous vein, particularly in surgically corrected patients, supporting individualized, substrate-guided ablation in this population.

**CLINICAL PERSPECTIVE:** *What Is Known:*

- Atrial fibrillation has been reported in adults with anomalous pulmonary venous return (APVR), but data on catheter ablation outcomes in this population are limited to case reports and small series.

*What the Study Adds:*

- In the largest reported cohort of patients with APVR undergoing catheter ablation for atrial fibrillation, pulmonary vein isolation achieved acute success in all patients despite substantial comorbidity burden and anatomic complexity.
- Non-pulmonary-vein sites of electrical activity were more common among patients with prior surgical correction, and postsurgical scar and epicardial venoatrial connections were also arrhythmogenic, indicating that the arrhythmogenic substrate in APVR often extends beyond the anomalous vein itself.
- These findings support individualized, substrate-guided mapping in patients with APVR, particularly those with a surgical history, and suggest a role for pulsed-field ablation near critical structures such as the phrenic nerve.

## INTRODUCTION

Anomalous pulmonary venous return (APVR) is a congenital abnormality in which one or more pulmonary veins drain outside the left atrium, most commonly into the right atrium or systemic venous circulation (1). Atrial fibrillation (AF) has been observed in adults with APVR and may be related to electrical activity from the anomalous pulmonary veins themselves (1, 2). Catheter ablation of AF with pulmonary vein isolation (PVI) is well-established, as pulmonary veins are a common source of arrhythmogenic triggers (3). However, the location of the anomalous pulmonary veins in patients with APVR can make PVI more challenging because these veins often lie close to the phrenic nerve or sinus node (1, 4).

Few reports describe PVI in patients with APVR, and there is no standard approach to whether the anomalous veins should be mapped and isolated (1, 4, 5). In a five-patient series of congenital anomalous cardiac veins undergoing pulsed-field ablation, anomalous pulmonary veins were successfully isolated without procedural complications, although follow-up was short (4). A review of 24 previously reported APVR cases found AF recurrence in five patients, none of whom underwent isolation of the anomalous pulmonary vein. In contrast, no recurrence was reported among patients with pulmonary vein potentials who underwent isolation (1). Because these findings were based on previously published cases, it remains unclear whether isolation affects long-term atrial arrhythmia recurrence.

In this case series, we describe procedural and long-term outcomes in patients with APVR and varying pulmonary venous anatomy who underwent PVI for atrial fibrillation.

## METHODS

### Study design and population

This is a retrospective, multicenter case series including adult patients (>18 years, non-pregnant) with confirmed anomalous pulmonary venous return (APVR), surgically corrected or uncorrected, who underwent catheter ablation for pulmonary vein isolation (PVI) at three tertiary academic centers in the United States between July 2003 and May 2025. Patients were identified from an institutional Epic-based database. The index procedure was defined as the first documented PVI performed for APVR-associated atrial fibrillation. Patients with unconfirmed or ruled-out APVR were excluded, as were patients with confirmed APVR who underwent ablation for other arrhythmias (atrioventricular nodal reentrant tachycardia, atrial flutter, focal atrial tachycardia, ventricular tachycardia, or premature ventricular contractions) without PVI, or who underwent only surgical ablation (e.g., Maze procedure) without a subsequent catheter-based PVI. Figure 1 was created with BioRender.com and is published with permission.

**Figure 1.**
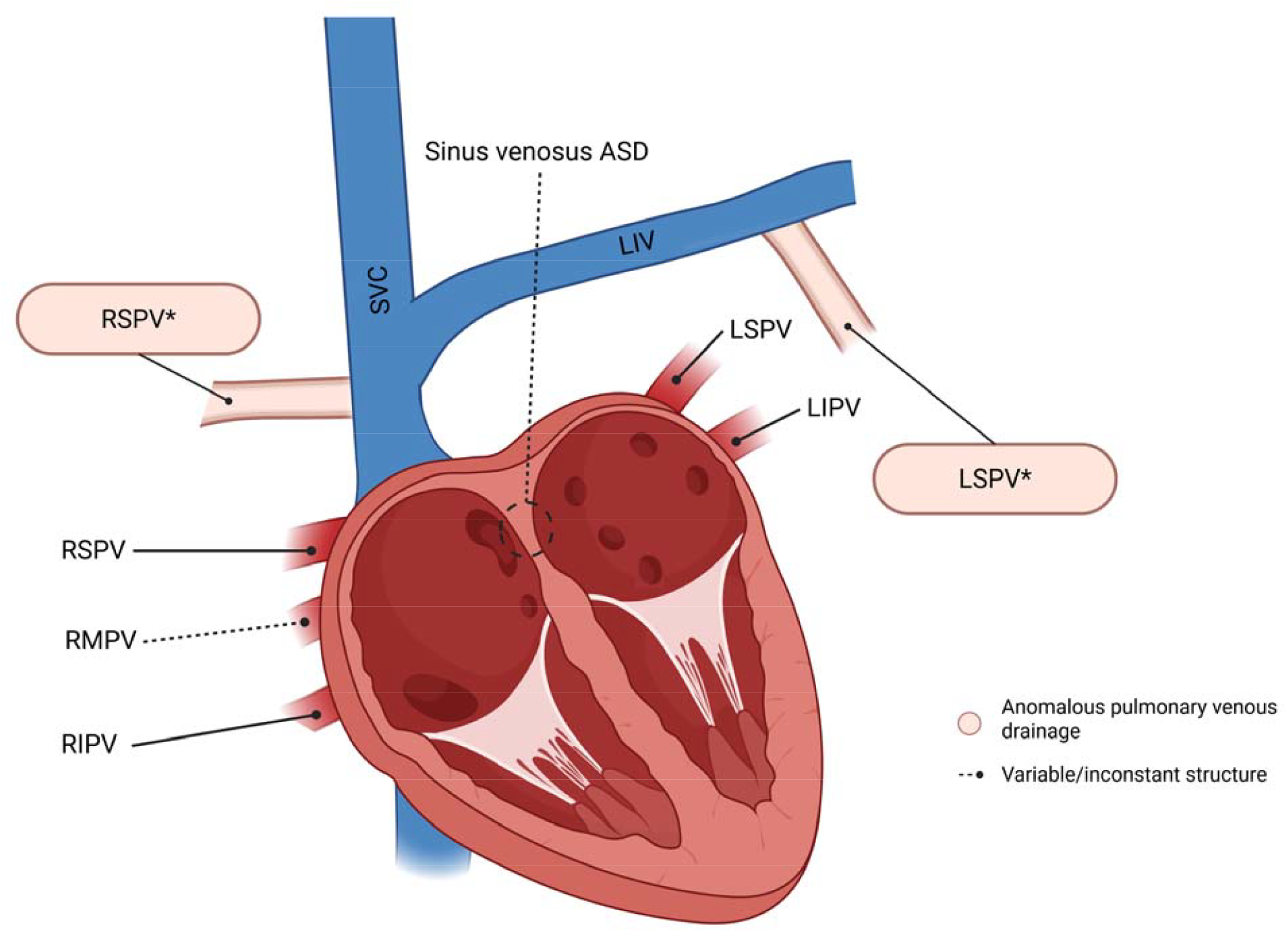
Representative patterns of pulmonary venous anomaly encountered in this cohort. Normal right- and left-sided pulmonary veins (RSPV, RMPV, RIPV, LSPV, LIPV) drain into the left atrium; the right middle pulmonary vein (RMPV) is a variable/inconstant structure present in only a subset of individuals. Asterisks denote representative anomalous drainage patterns observed in the cohort: right pulmonary veins draining into the SVC (RSPV*) and left pulmonary veins draining into the left innominate vein (LSPV*). A concomitant sinus venosus ASD (dashed outline) was also observed. Total APVR, in which all pulmonary veins drain anomalously, was present in 2 patients and is not depicted. SVC indicates superior vena cava; LIV, left innominate vein; ASD, atrial septal defect. Created with BioRender.com.

### Endpoints and definitions

Recurrence was defined as any symptomatic or asymptomatic atrial arrhythmia (AF, atrial flutter, or atrial tachycardia) lasting ≥30 seconds, documented after a 90-day blanking period. Early recurrence was defined as any atrial arrhythmia within the 90-day blanking period and was further classified as immediate (0-30 days) or delayed (31-90 days). The need for reablation was recorded. Periprocedural complications were defined as any adverse event occurring within 30 days of the index procedure and classified by timing as early (0-48 hours) or late (48 hours to 30 days).

### Follow-up

Follow-up was assessed through any arrhythmia-related visit or call (electrophysiology, cardiology, internal medicine, or family medicine) documented in the electronic health record. Device interrogation data and external ECG monitoring results were collected.

The study was approved by the Institutional Review Board of our institution (protocol no. 25-012938), which granted exempt status. The data supporting the findings of this study are available from the corresponding author upon reasonable request.

### Data analysis

Continuous variables are reported as mean ± SD or median (IQR), depending on distribution; categorical variables as frequency (%). Given the small sample size and low event rate, no formal statistical comparisons between subgroups were performed; findings are presented descriptively. Analyses were performed and figures generated using R (version 4.6.0; R Foundation for Statistical Computing, Vienna, Austria).

## RESULTS

Fourteen patients met inclusion criteria (Table 1). Mean age at the index procedure was 56 ± 13 years, and 11 (79%) were male. Median BMI was 27 kg/m^2^ (IQR, 26-31; reported as median due to a high-BMI outlier). Paroxysmal AF was the predominant presentation (9/14, 64%). Mean CHA2DS2-VA score was 2 ± 2. CHA2DS2-VA (rather than CHA2DS2-VASc) was used given growing evidence supporting sex-independent stroke risk stratification, as adopted in the 2024 ESC guidelines for the management of atrial fibrillation (6). Comorbidities included heart failure (9, 64%), hypertension (6, 43%), coronary artery disease (6, 43%), obstructive sleep apnea (6, 43%), diabetes mellitus (3, 21%), chronic kidney disease (2, 14%), and peripheral artery or pulmonary disease (1 each, 7.1%). Mean LVEF was 55% ± 10%, mean LAVI was 37 ± 10 mL/m^2^, and mean RVSP was 33 ± 8 mmHg; moderate-severe valvular disease was present in 6 patients (43%). Baseline rhythm was sinus in 9 (64%), atrial fibrillation in 1 (7.1%), atrial flutter in 2 (14%), and paced in 2 (14%). Most patients had partial APVR (12, 86%), and 2 (14%) had total APVR. Eight patients (57%) had undergone prior surgical correction of their congenital venous anomaly. Anatomic distribution of the anomalous drainage is shown in Figure 1: right pulmonary veins draining into the SVC (RSPV*) in 6 patients (42.9%) and left pulmonary veins draining into the left innominate vein (LSPV*) in 2 patients (14.3%). The specific native drainage pattern could not be determined in the remaining 4 patients, all with prior surgical correction, as contemporary imaging reflected the postoperative venous pathway rather than the native anomaly. A concomitant sinus venosus atrial septal defect was present in 5 patients (35.7%), including 2 of the 6 RSPV* patients.

**Table 1.**
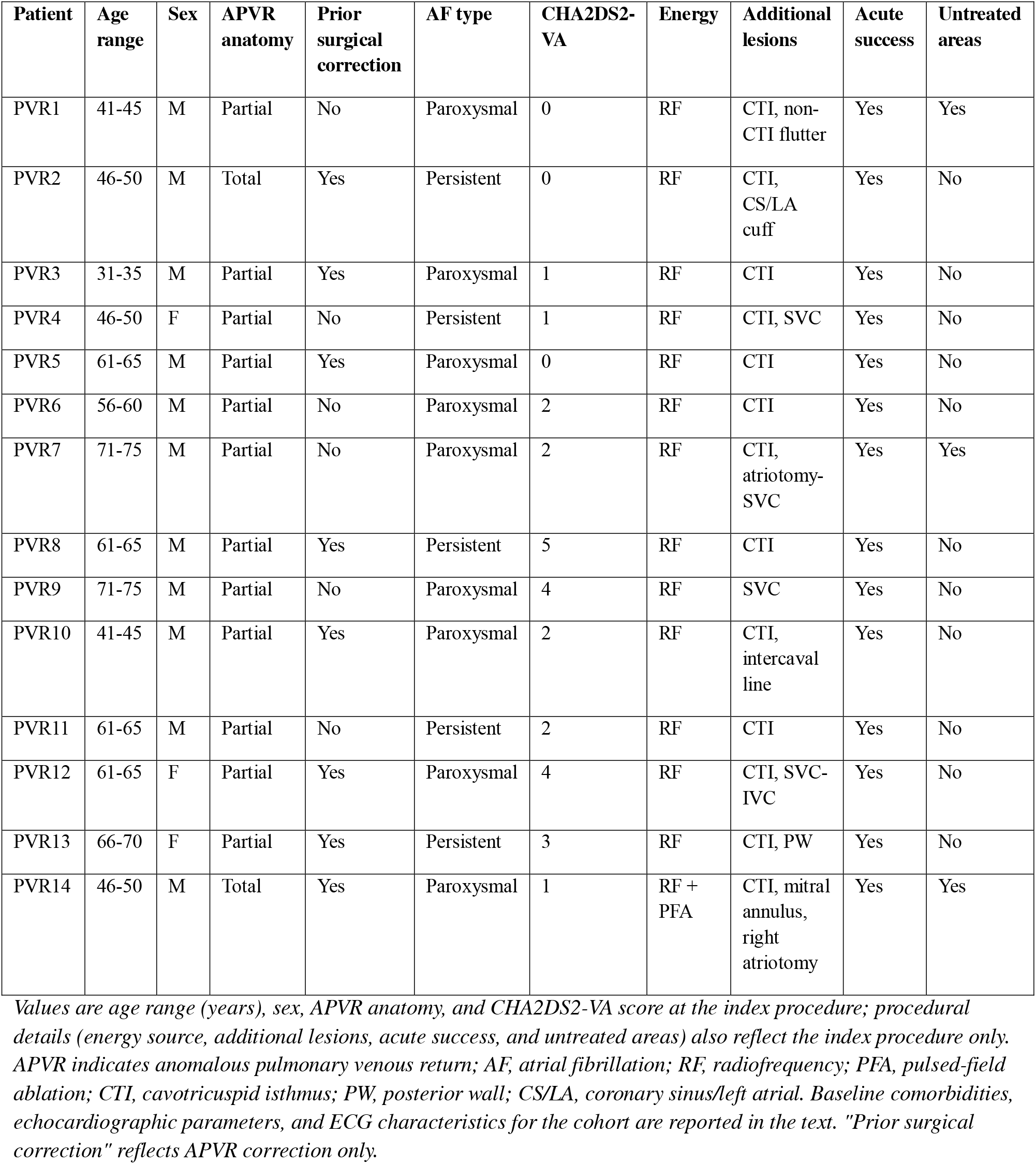
Patient Characteristics and Procedural Findings.

| Patient | Age range | Sex | APVR anatomy | Prior surgical correction | AF type | CHA2DS2-VA | Energy | Additional lesions | Acute success | Untreated areas |
| --- | --- | --- | --- | --- | --- | --- | --- | --- | --- | --- |
| PVR1 | 41-45 | M | Partial | No | Paroxysmal | 0 | RF | CTI, non-CTI flutter | Yes | Yes |
| PVR2 | 46-50 | M | Total | Yes | Persistent | 0 | RF | CTI, CS/LA cuff | Yes | No |
| PVR3 | 31-35 | M | Partial | Yes | Paroxysmal | 1 | RF | CTI | Yes | No |
| PVR4 | 46-50 | F | Partial | No | Persistent | 1 | RF | CTI, SVC | Yes | No |
| PVR5 | 61-65 | M | Partial | Yes | Paroxysmal | 0 | RF | CTI | Yes | No |
| PVR6 | 56-60 | M | Partial | No | Paroxysmal | 2 | RF | CTI | Yes | No |
| PVR7 | 71-75 | M | Partial | No | Paroxysmal | 2 | RF | CTI, atriotomy-SVC | Yes | Yes |
| PVR8 | 61-65 | M | Partial | Yes | Persistent | 5 | RF | CTI | Yes | No |
| PVR9 | 71-75 | M | Partial | No | Paroxysmal | 4 | RF | SVC | Yes | No |
| PVR10 | 41-45 | M | Partial | Yes | Paroxysmal | 2 | RF | CTI, intercaval line | Yes | No |
| PVR11 | 61-65 | M | Partial | No | Persistent | 2 | RF | CTI | Yes | No |
| PVR12 | 61-65 | F | Partial | Yes | Paroxysmal | 4 | RF | CTI, SVC-IVC | Yes | No |
| PVR13 | 66-70 | F | Partial | Yes | Persistent | 3 | RF | CTI, PW | Yes | No |
| PVR14 | 46-50 | M | Total | Yes | Paroxysmal | 1 | RF + PFA | CTI, mitral annulus, right atriotomy | Yes | Yes |

Values are age range (years), sex, APVR anatomy, and CHA2DS2-VA score at the index procedure; procedural details (energy source, additional lesions, acute success, and untreated areas) also reflect the index procedure only. APVR indicates anomalous pulmonary venous return; AF, atrial fibrillation; RF, radiofrequency; PFA, pulsed-field ablation; CTI, cavotricuspid isthmus; PW, posterior wall; CS/LA, coronary sinus/left atrial. Baseline comorbidities, echocardiographic parameters, and ECG characteristics for the cohort are reported in the text. "Prior surgical correction" reflects APVR correction only.

### Procedural findings

Patient-level procedural details are included in Table 1. Radiofrequency ablation was used in all patients at the index procedure, combined with pulsed-field ablation in 1 patient (7.1%, dual-energy RF/PFA catheter). The ablation catheter was documented in 6 patients (ThermoCool in 4, QDOT Micro in 1, Sphere-9 in 1). Mean procedure time was 351 ± 108 minutes, and median fluoroscopy time was 24.5 minutes (IQR, 18.6-41.6; reported as median due to one outlier case of 93 minutes). In 3 patients (21%), the index procedure in this series was a repeat ablation following an earlier ablation.

Pulmonary vein isolation was achieved in all patients. Additional lesion sets included cavotricuspid isthmus ablation in 13 patients (92.9%), posterior wall isolation in 1 (7.1%), and other adjunctive lesion sets in 8 (57.1%).

On electroanatomic mapping (Figure 2), the distribution of documented electrical activity varied across mapping sites, with no consistent difference between patients with (n=7) and without (n=6) prior surgical correction (n=13; mapping data were unavailable in 1 patient). Non-pulmonary-vein (“Other”) sources of activity were more common among patients with prior surgical correction (43%) than among those without (17%).

**Figure 2.**
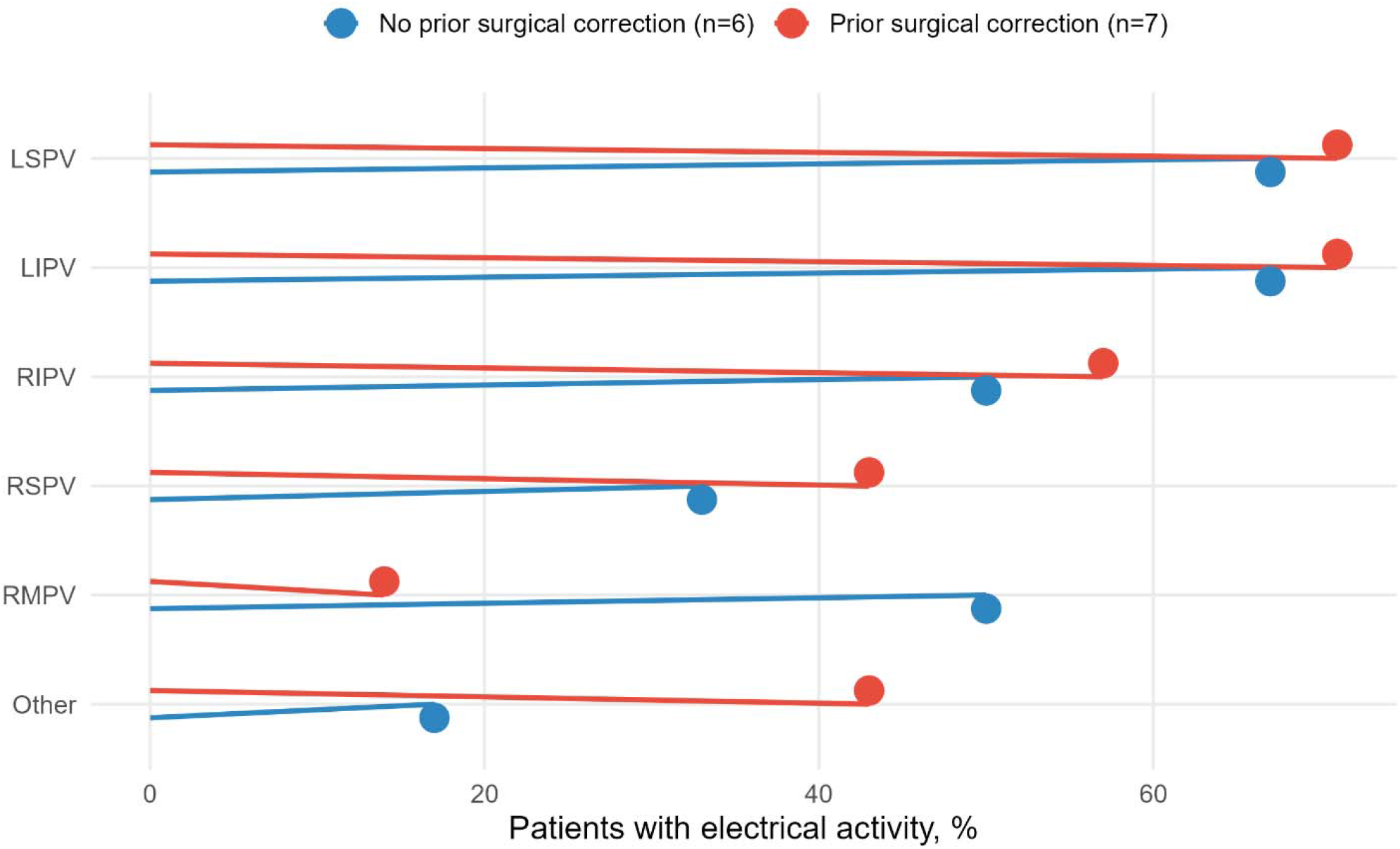
Electrical activation by pulmonary vein mapping site, stratified by prior surgical correction (n=13). Percentage of patients with documented electrical activity at each mapping site during electroanatomic mapping, compared between patients without (n=6) and with (n=7) prior surgical correction of their congenital anomaly. One patient (7%) was excluded from this analysis due to unavailable mapping data. LSPV indicates left superior pulmonary vein; LIPV, left inferior pulmonary vein; RIPV, right inferior pulmonary vein; RSPV, right superior pulmonary vein; and RMPV, right middle pulmonary vein.

Untreated areas with documented electrical activity remained after the index procedure in 3 patients (21%). In 1, the right middle pulmonary vein remained connected to the right atrium and, together with the adjacent SVC, was left unablated because of phrenic nerve capture. In a second, no ablation was performed at the SVC/right atrial junction for the same reason. In a third, the superior portion of a surgical atriotomy scar extending to the SVC was not ablated because of proximity to the sinus node; this patient later developed postprocedural PVCs and ventricular tachycardia.

### Periprocedural complications

Periprocedural complications occurred in 3 patients (21%), all within 48 hours of the procedure. These included access-site-related bleeding (managed conservatively, with stable hemoglobin) in 2 patients, and postprocedural PVCs and ventricular tachycardia necessitating an extended hospital stay and initiation of oral mexiletine and amiodarone in 1 patient. This patient had no early or late recurrence of atrial arrhythmias and no other complication: pulmonary vein isolation remained durable at a repeat procedure 9 months later for ventricular arrhythmia.

### Outcomes

Nine patients (64%) were discharged on antiarrhythmic or beta-blocker therapy (metoprolol in 4, amiodarone in 2, and dofetilide, dronedarone, and mexiletine in 1 each), and 9 were discharged on a direct oral anticoagulant and 5 on warfarin. Median follow-up was 3.4 years (IQR, 2.0-9.6; range, 0.9-15.4). Early recurrence within the 90-day blanking period was confirmed in 2 patients (14%): one immediate (0-30 days) and one delayed (31-90 days). Long-term recurrence occurred in 7 patients (50%): 5 atrial fibrillation and 2 atrial flutter. Both patients with early recurrence within the blanking period ultimately required reablation (Figure 3): one for a non-pulmonary-vein-dependent mitral isthmus flutter, successfully ablated with radiofrequency energy despite durable isolation of the common pulmonary vein trunk; and one for roof-dependent flutter with reconnection along the left-sided veins, in whom ablation was switched to pulsed-field energy because of esophageal proximity, with acute termination of flutter and re-isolation achieved.

**Figure 3.**
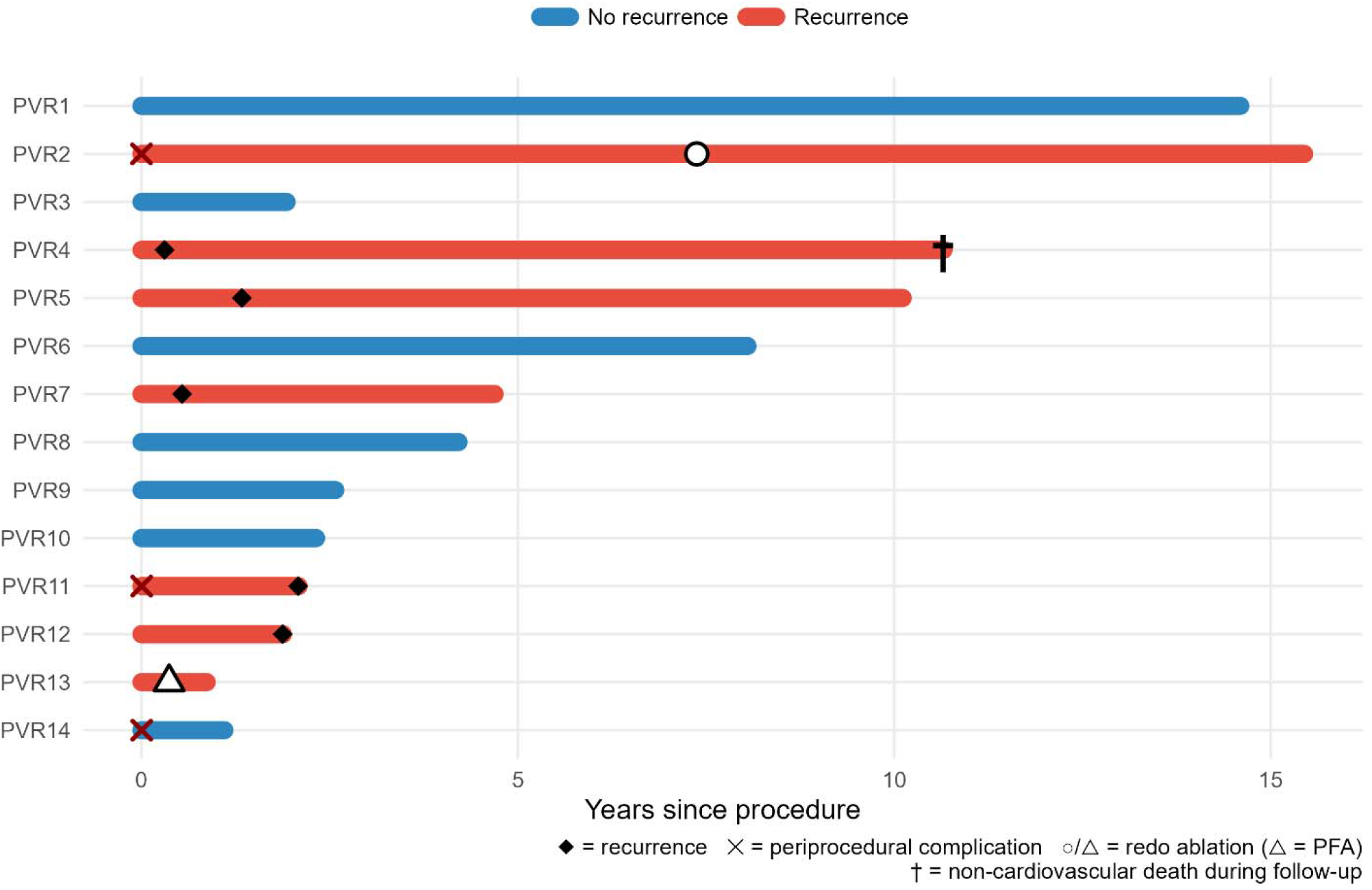
Follow-up timeline (n=14). Bars represent duration of follow-up from the index procedure, color-coded by recurrence status. Diamonds indicate the time of arrhythmia recurrence; X marks indicate periprocedural complications; circles and triangles indicate redo ablation procedures (triangle denotes use of pulsed-field ablation); and a dagger (†) indicates non-cardiovascular death during follow-up.

Freedom from atrial arrhythmia recurrence, estimated by Kaplan-Meier analysis (Figure 4), was 78.6% (95% CI, 59.8-100%) at 1 year and 53.9% (95% CI, 32.3-89.9%) at 5 years. One non-cardiovascular death occurred during follow-up.

**Figure 4.**
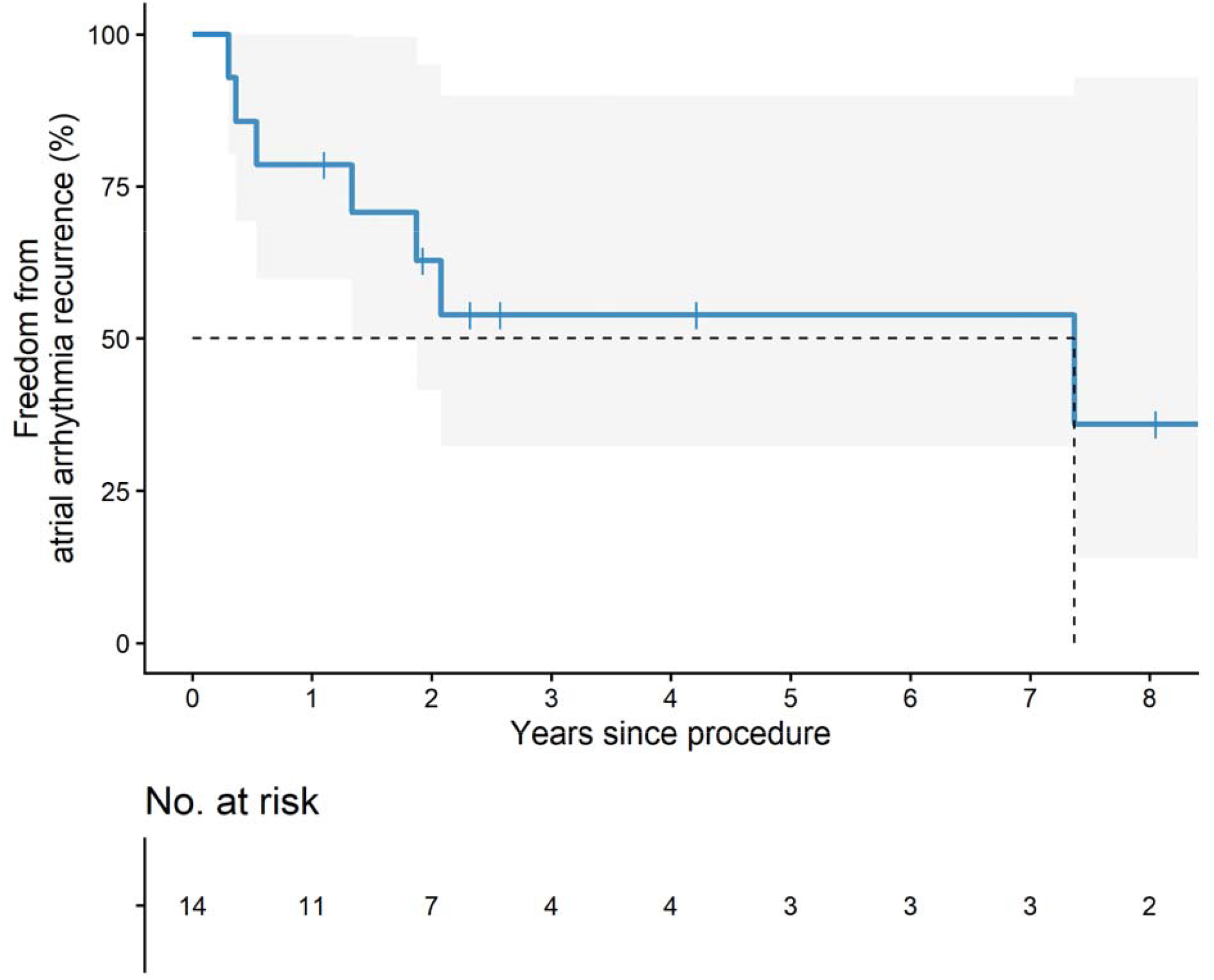
Kaplan-Meier estimate of cumulative freedom from atrial arrhythmia recurrence after catheter ablation in patients with anomalous pulmonary venous return (n=14). Freedom from atrial arrhythmia recurrence was 78.6% (95% CI, 59.8-100%) at 1 year and 53.9% (95% CI, 32.3-89.9%) at 5 years. Shaded area represents the 95% confidence interval; vertical tick marks indicate censored patients. The curve is truncated at 8 years, beyond which fewer than two patients remained at risk.

## DISCUSSION

To our knowledge, this is the largest cohort of patients with APVR undergoing catheter ablation for atrial fibrillation reported to date, exceeding the previously largest reported series of 9 patients (5). Procedures were long, with a mean procedure time of 351 ± 108 minutes, and most patients required ablation beyond PVI, including cavotricuspid isthmus lines in 92.9% and adjunctive lesion sets in 57.1%. Despite this complexity, acute success was achieved in all patients, although residual areas with documented electrical activity were intentionally left untreated in 3 (21%). Non-pulmonary-vein activation was also more common among patients with prior surgical correction of their venous anomaly than among those without (43% vs 17%), suggesting a substrate that in this subgroup may be more postsurgical than purely venous in origin. Freedom from atrial arrhythmia recurrence at 5 years (53.9%) was similar to the pooled 5-year single-procedure success rate reported for AF ablation in the general population with AAD (7), suggesting that the anatomic and comorbidity burden of APVR did not reduce longterm durability compared with standard AF ablation.

Our arrhythmia-free survival compares favorably with reported outcomes in broader ACHD populations undergoing AF ablation, where 1-year freedom from recurrence has generally ranged from 45% to 63%, including a cohort of 145 patients with mixed CHD lesions (8), and where 5-year freedom was as low as 22% in one long-term series (9). This may reflect the simpler anatomic substrate of isolated APVR compared with the more complex lesions, such as Fontan palliation or transposition of the great arteries, that predominate in those cohorts, rather than any difference in ablation technique. Within the APVR-specific literature, our 1-year freedom from recurrence (78.6%) is lower than the 89% reported by Demarchi et al. (5), though both series are small and this difference should be interpreted cautiously.

Pulmonary veins are the dominant source of atrial ectopic triggers (3), but our findings suggest that this does not fully apply in APVR. This fits with the broader ACHD literature, where atriotomy scars, baffles, and suture lines are recognized as an arrhythmogenic substrate distinct from pulmonary vein triggers, and where extra-pulmonary triggers challenge ablation strategies built around PVI alone (10). In one of our patients without prior surgical correction of the anomalous vein, mapping identified an epicardial connection between the right middle pulmonary vein and the right atrium; both this connection and the adjacent SVC were intentionally left unablated because of phrenic nerve capture on high-output pacing. This mirrors the mechanism described by Bhalla et al. (2) and suggests that such epicardial venoatrial connections are not necessarily tied to prior cardiac surgery. In a second patient, the superior portion of a surgical atriotomy scar extending toward the SVC was left unablated because of its proximity to the sinus node, illustrating how postsurgical scar itself can become a distinct ablation target, separate from the anomalous vein. Together with the higher rate of non-venous activation seen among surgically corrected patients on mapping, these cases support a lower threshold for broader substrate mapping in patients with a surgical history, rather than focusing on the anomalous vein alone.

In each of the three patients with untreated areas of documented electrical activity, ablation was withheld deliberately, usually because of proximity to the phrenic nerve or sinus node, rather than because of a procedural limitation. The patient with the RMPV-right atrial connection described above had long-term, rather than early, arrhythmia recurrence originating from that same untreated area, managed without reablation. This pattern echoes Wei et al.’s review of 24 previously published APVR cases, in which recurrence occurred only among patients whose anomalous vein or pulmonary vein potentials were not isolated, while none occurred among those who underwent isolation (1). Our numbers are too small to draw firm conclusions, but the consistency across independent series supports ablating sites of documented electrical activity whenever this can be done safely.

Proximity to the phrenic nerve or sinus node was the reason ablation was withheld in our cohort, and this constraint is where pulsed-field ablation (PFA) may be most useful. Periprocedural complications were infrequent, and phrenic nerve injury, the concern most often raised in this anatomic context (1, 4), did not occur, in part because ablation was withheld at sites of phrenic capture. The nonthermal mechanism of PFA may allow some of these sites to be targeted safely: it was applied without complication in 5 patients with congenital venous anomalies in one series (4), and in our cohort it was chosen for a redo procedure because esophageal proximity had limited radiofrequency delivery at the index ablation. With growing experience, PFA may allow ablation of substrate that currently must be left untreated.

## LIMITATIONS

This study has several limitations. The design was retrospective and multicenter, which precludes causal inference. The small sample size limited analyses to descriptive comparisons without formal statistical testing between subgroups. Recording of some baseline and echocardiographic variables was incomplete, and follow-up was heterogeneous, spanning a 22-year period (2003-2025) across multiple ablation technology eras, from radiofrequency-only practice to the recent introduction of pulsed-field ablation. The referral-based tertiary/quaternary population may limit generalizability, and mapping and ablation strategies were guided by operator judgment rather than a standardized protocol. Given the rarity of APVR, larger prospective, multicenter studies are needed to validate these findings, formally test whether prior surgical correction predicts a distinct arrhythmogenic substrate, and define the long-term role of pulsed-field ablation in this population.

## CONCLUSIONS

In this largest reported cohort of patients with APVR undergoing catheter ablation for atrial fibrillation, PVI achieved acute success in all patients despite a high burden of comorbidities and anatomic complexity. Non-pulmonary-vein sites of electrical activity were more common among surgically corrected patients, and postsurgical scar and epicardial venoatrial connections were also arrhythmogenic, suggesting that the arrhythmogenic substrate in APVR often extends beyond the anomalous vein itself, particularly after prior cardiac surgery. Complications were infrequent, and pulsed-field ablation may improve the safety of ablation near critical structures such as the phrenic nerve. These findings support individualized, substrate-guided mapping in this population and warrant validation in larger, prospective studies.

## Data Availability

The data supporting the findings of this study are available from the corresponding author upon reasonable request.

## ACKNOWLEDGMENTS

None.

## AUTHOR CONTRIBUTIONS

All authors contributed equally to the concept and design of the study, data collection and analysis, drafting and critical revision of the manuscript, and approval of the final version submitted.

